# Study COVID-19 Severity of Patients Admitted to Emergency Room (ER) with Chest X-ray Images

**DOI:** 10.1101/2022.12.25.22283942

**Authors:** Jonathan Stubblefield, Christopher Saldivar, Anna De Feria, James Riddle, Abhijit Shivkumar, Jason Causey, Jake Qualls, Jennifer Fowler, Xiuzhen Huang

**Author notes:** **Data statement.** The clinical data and chest x-ray image data for this study were collected and prepared by the residents and researchers of the Joint Translational Research Lab of Arkansas State University (A-State) and St. Bernards Medical Center (SBMC) Internal Medicine Residency Program. As data collection is on-going for the project stage-II of clinical testing, raw data is not currently available for data sharing to the public. **Ethics.** This study was approved by the St. Bernards Medical Center’s Institutional Review Board (IRB).

## Abstract

We have conducted a study of the COVID-19 severity with the chest x-ray images, a private dataset collected from our collaborator St Bernards Medical Center. The dataset is comprised of chest x-ray images from 1,550 patients who were admitted to emergency room (ER) and were all tested positive for COVID-19. Our study is focused on the following two questions: (1) To predict patients hospital staying duration, based on the chest x-ray image which was taken when the patient was admitted to the ER. The length of stay ranged from zero hours to 95 days in the hospital and followed a power law distribution. Based on our testing results, it is hard for the prediction models to detect strong signal from the chest x-ray images. No model was able to perform better than a trivial most-frequent classifier. However, each model was able to outperform the most-frequent classifier when the data was split evenly into four categories. This would suggest that there is signal in the images, and the performance may be further improved by the addition of clinical features as well as increasing the training set. (2) To predict if a patient is COVID-19 positive or not with the chest x-ray image. We also tested the generalizability of training a prediction model on chest x-ray images from one hospital and then testing the model on images captures from other sites. With our private dataset and the COVIDx dataset, the prediction model can achieve a high accuracy of 95.9%. However, for our hold-one-out study of the generalizability of the models trained on chest x-rays, we found that the model performance suffers due to a significant reduction in training samples of any class.

## 1. Background and Introduction

The global pandemic caused by Sars-CoV-2, also known as COVID-19 has had a significant global impact [McIntosh2020], not only resulting in 6,678,877 deaths at the time of this writing [Link], but also has lasting morbidity in certain survivors, to say nothing of the economic consequences. Not all cases of Sars-CoV-2 were equally severe. It is difficult to state with certainty the percentage of infected patients who never developed symptoms as these patients may not even present for medical attention. Additionally, different studies examining this issue assessed for different symptoms and thus would be expected to classify patients differently. A meta-analysis of studies estimates that 33 percent of unvaccinated, infected patients could be expected to be asymptomatic.

Among symptomatic patients, severity still varies. An early report from the Chinese Center for Disease Control and Prevention [Link1] provides some insight. Typically, 81% of patients have mild disease. 14% of patients have severe disease, defined as the presence of dyspnea, hypoxia, or greater than 50% of lung involvement on imaging. 5% of patients had critical disease, defined as respiratory failure, shock, or multiorgan dysfunction. The overall case fatality rate was 2.3%.

A similar report by the United States Centers for Disease Control and Prevention [Link2] showed that 14% of patient were admitted to the hospital. Another 2% of patients were admitted to the intensive care unit (ICU). The case fatality rate was 5%.

It would be particularly valuable for physicians to be able to predict the severity of Sars-CoV-2 early in the patient’s presentation. Knowing ahead of time if the patient is likely to be admitted to the ICU, require a ventilator, or die, or knowing the number of hospital days a patient will likely require, can guide patient care as well as the allocation of hospital resources.

Chest x-rays are frequently obtained early during a hospital stay for initial patient evaluation. When chest x-rays show signs of infection, Sars-CoV-2 typically appears as an atypical pneumonia with consolidation, ground glass opacities, and diffuse involvement across the lower lungs bilaterally and peripherally. However, the imaging findings are usually non-specific and not informative.

We have been collaborating with the Internal Medicine Residency Program of St Bernards Medical Center through the Joint Translational Research Lab for imaging and clinical data collection of patients admitted to ER. In a previous related work [Stubblefield], we evaluated emergency room (ER) patient classification for cardiac and infection causes with clinical data and chest X-ray image data. With this study we collaborated with St Bernards Medical Center Internal Medicine Residency Program to collect 1550 COVID-19 patients chest x-ray images and conducted the severity analysis of these patients with chest x-ray images. Our study is focused on the following two questions: (1) To predict patients hospital staying duration, based on the chest x-ray image which was taken when the patient was admitted to the ER. (2) To predict if a patient is COVID-19 positive or not with the chest x-ray image. We also tested the generalizability of training a prediction model on chest x-ray images from one hospital and then testing the model on images captures from other sites.

## 2. Predict hospital staying duration based on chest x-ray image

### Chest x-ray image dataset from St. Berbards Medical Center

The imaging dataset is comprised of chest x-rays from 1,550 patients who all tested positive for COVID-19. We have additional more images which were collected; however, these images were excluded from this study, since they did not have associated hospital duration data. The data acquisition protocol was that each sample x-ray was taken from the first set of x-rays the patient received after being admitted. The dataset also included the duration of the patient’s stay in the hospital. The length of stay ranged from zero hours to 95 days in the hospital and followed a power law distribution. There were few patients on the upper end which presented problems with the models. This dataset will be supplemented with clinical features for our planned future study. The expectation is that this will assist in increasing the data available for patients with longer term hospital stays and improve performance. The TensorFlow image augmentation generator was used to enrich the dataset. The images were augmented by shearing, rotating, horizontal flipping, height and width shifting, brightness, and zoom.

### Models to predict ER patients’ hospital staying duration

The dataset was used to predict the duration based on one of four bins. A pre-trained DenseNet model using ImageNet weights and another using CheXNet weights were used. Given the difficulties presented by the data’s distribution I chose to perform a classification task on this dataset. The patients were split into four categories based on their length of stay. The research task was to predict which category each patient belonged to based solely on their chest x-ray. These four categories were determined by using the Fisher-Jenks algorithm for finding “natural breaks” in the data. Fisher-Jenks clusters the data by minimizing the variance between members of the cluster and maximizing the variance between clusters. Four clusters were chosen because any more simply split the patients with longer durations and the number of patients in these categories was too low.

The four categories were split at 8.21 days, 18.83 days, 42.45 days, and 95.79 days. 1,079 patients were in the first category, 368 were in the second, 148 were in the third, and 8 were in the last.

Two pretrained models were used for the classification task: ImageNet and CheXNet. ImageNet is a 121-layer DenseNet architecture that was pre-trained on the ImageNet dataset, consisting of 1.4 million images representing 1,000 categories. CheXNet is also a 121-layer DenseNet architecture; however, it was trained using NIH ChestX-ray14 dataset, consisting of 100,000 frontal x-rays This was done to compare the improvement of using a model pretrained on a domain relevant dataset relative to the same architecture pretrained on a different domain.

The final output layer for the CheXNet model was removed and the last fully connected layer with 1,024 features were used during training. The rationale was that this model was used to extract features from the x-ray images and this last layer encoded the relevant feature which were used for training. Another experiment was also conducted where the final prediction layer was used as a feature extractor rather than the last dense layer. The final 14 predicted probabilities were used as extracted features and used as input to the models for training. This was done to compare the relative performance of using a reduced number of features for prediction.

Three non-deep learning models were also trained to compare the performance against the neural network. All the models were trained using 14 CheXNet predictions rather than the images directly. This was chosen so that a more direct comparison could be made between the models. The neural network would have had an unfair advantage over the other models since the training set was enriched using augmented images as previously discussed. These images were fed directly into the CheXNet model, and the prediction vectors were saved for 30 epochs worth of training. This dataset was then used to train all the models so that they would all have access to the exact same training and validation samples.

XGBoost was trained using the following parameters: “eta”: 0.3, “max_depth”: 2, “objective”: “multi:softmax”, “max_depth”: 6. We also used k-Nearest Neighbors and logistic regression models to compare against. Knn was trained using k=5 and logistic regression was training using the settings max_iter=1000 and class_weight. Each model was also evaluated using 5-fold cross-validation and the same folds were used for each model. Each model was also trained on both categories derived from the Fischer-Jenks algorithm as well as the categories where the data was split evenly between four classes.

### The performance of different prediction models compared with the baseline classifier

A most-frequent classifier was tested prior to evaluating the trained models. This classifier simply chooses the most frequent class in the training set regardless of the inputs. Bin 1 had the most class samples at 1,027 and so the dummy classifier had an accuracy of 66 %. This represents the accuracy one should expect by simply guessing and serves as a baseline to evaluate the models. The ImageNet model had 64.14% (SD=4.18%) accuracy after training and the CheXNet model had a 68.39% accuracy (SD=3.05%). The ChexNet model was then retrained using class weights and fine-tuning. This resulted in an increase to 71.61% accuracy and a weighted f1 score of 69.34%. The CheXNet model was able to slightly outperform the most-frequent classifier with this heavily imbalanced dataset. It performed poorly with groups 3 and 4. These groups represented 9.5% and 0.5% of the data in the training set respectively.

The experiments were executed again using 5-fold cross validation and the ChexNet derived model was no longer able to outperform a most-frequent classifier. The neural-network and xgboost models performed the same as the dummy classifier.

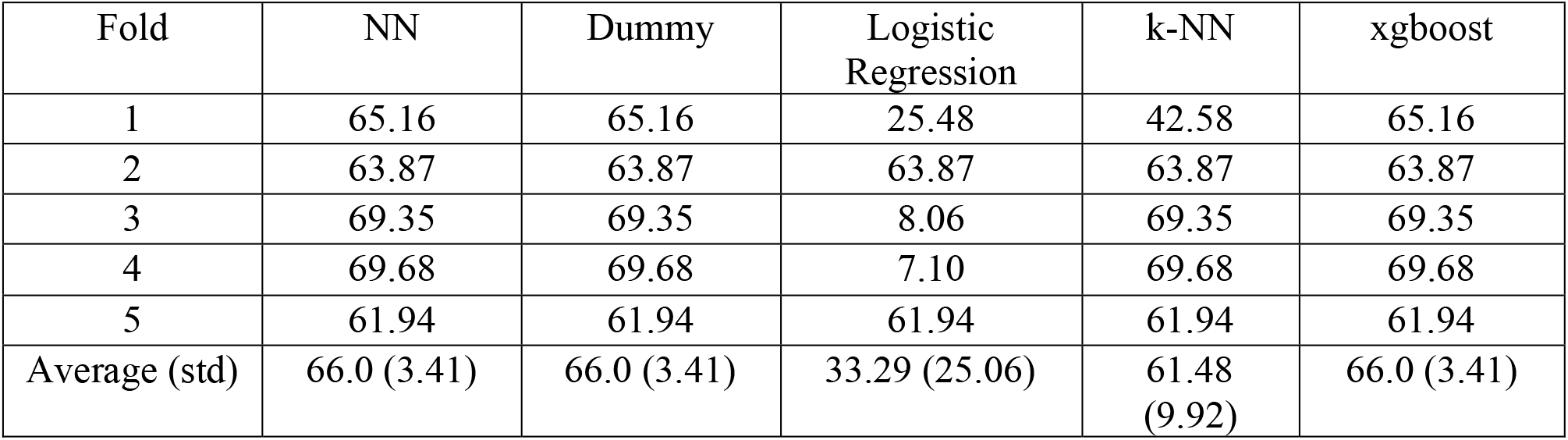

| Fold | NN | Dummy | Logistic Regression | k-NN | xgboost |
| --- | --- | --- | --- | --- | --- |
| 1 | 65.16 | 65.16 | 25.48 | 42.58 | 65.16 |
| 2 | 63.87 | 63.87 | 63.87 | 63.87 | 63.87 |
| 3 | 69.35 | 69.35 | 8.06 | 69.35 | 69.35 |
| 4 | 69.68 | 69.68 | 7.10 | 69.68 | 69.68 |
| 5 | 61.94 | 61.94 | 61.94 | 61.94 | 61.94 |
| Average (std) | 66.0 (3.41) | 66.0 (3.41) | 33.29 (25.06) | 61.48 (9.92) | 66.0 (3.41) |

The data was then split evenly into four groups. The models were able all able to outperform the dummy classifier; however, performance was still poor. The neural network performed the best of all the models with an accuracy of 27.68% (STD 1.33%) and an f1-score of 24.89% (STD 2.89%).

| Fold | NN | Dummy | Logistic Regression | k-NN | xgboost |
| --- | --- | --- | --- | --- | --- |
| 1 | 25.48 | 21.29 | 31.61 | 30.32 | 21.29 |
| 2 | 29.03 | 21.29 | 21.61 | 22.90 | 26.12 |
| 3 | 27.10 | 25.16 | 23.87 | 26.45 | 23.55 |
| 4 | 29.03 | 20.00 | 22.90 | 21.29 | 28.39 |
| 5 | 27.74 | 22.58 | 26.77 | 29.68 | 25.16 |
| Average (std) | 27.68 (1.33) | 22.06 (1.75) | 25.35 (1.75) | 26.13 (3.58) | 24.9 (2.39) |

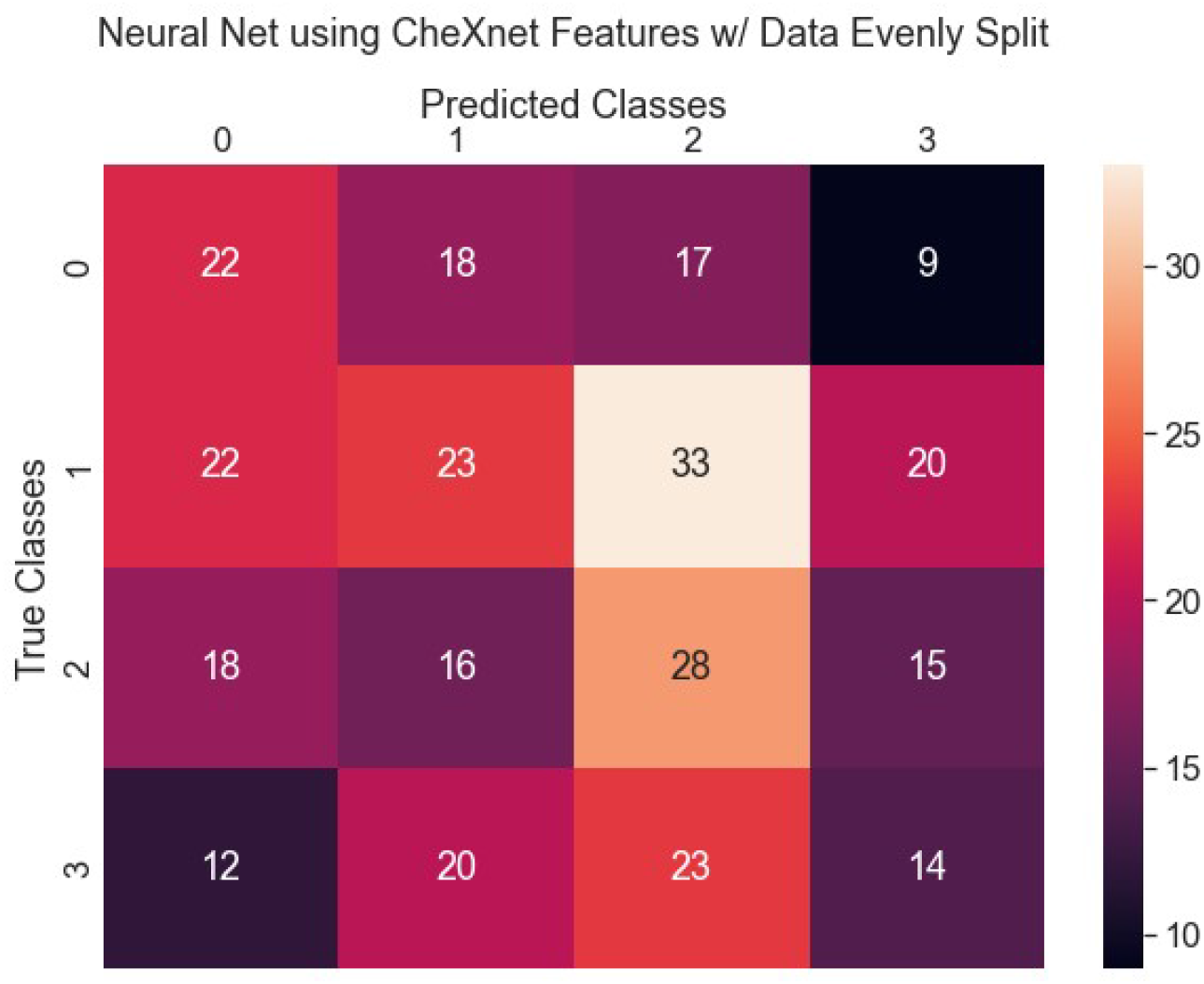

This confusion matrix shows how the neural network model predominately predicted patients in classes 1 and 2 followed closely by class 0. The model had the poorest performance with class 3 (the patients with the longest duration in the hospital). With the strongest performance in class 2. The accuracy for each class is as follows:

|  |  |
| --- | --- |
| Class 0 | 33.33% |
| Class 1 | 23.47% |
| Class 2 | 36.36% |
| Class 3 | 20.29% |

Based on the results of these tests the models were unable to detect strong signal from the chest x-rays they were trained on. No model was able to perform better than a trivial “dummy” most-frequent classifier when predicting the breaks derived using the Fisher-Jenks algorithm [Jenks, McMaster]. However, each model was able to outperform the most-frequent classifier when the data was split evenly into 4 categories. This would suggest that there is signal in the images; however, accuracy was still poor. This may be further improved by the addition of clinical features as well as increasing the training set. There were few samples of long hospital stays in the dataset. The mean duration was 8.4 days and a standard deviation of 8.29 days with 87.35% of the samples within one standard deviation of the mean. The models were not able to detect a discernible difference between these samples due to the high density of the samples and mis-predicted samples with long durations in the hospital.

## 3. Predict COVID-19 positive or negative with chest x-ray images

### Chest x-ray image dataset from St. Berbards Medical Center and COVIDx dataset

1,648 chest x-ray images from St. Bernard’s were used for this task. We were able to use more chest x-ray images from St. Bernard’s because we did not need to match the images with hospital durations, as was necessary in the previous task. We used the COVIDx dataset [Wang] to assist in training which provided an additional 16,344 images for a total of 17,992 images. It was necessary to use the additional dataset because the St. Bernard images only contained positive cases and simply to increase the size and diversity of the dataset.

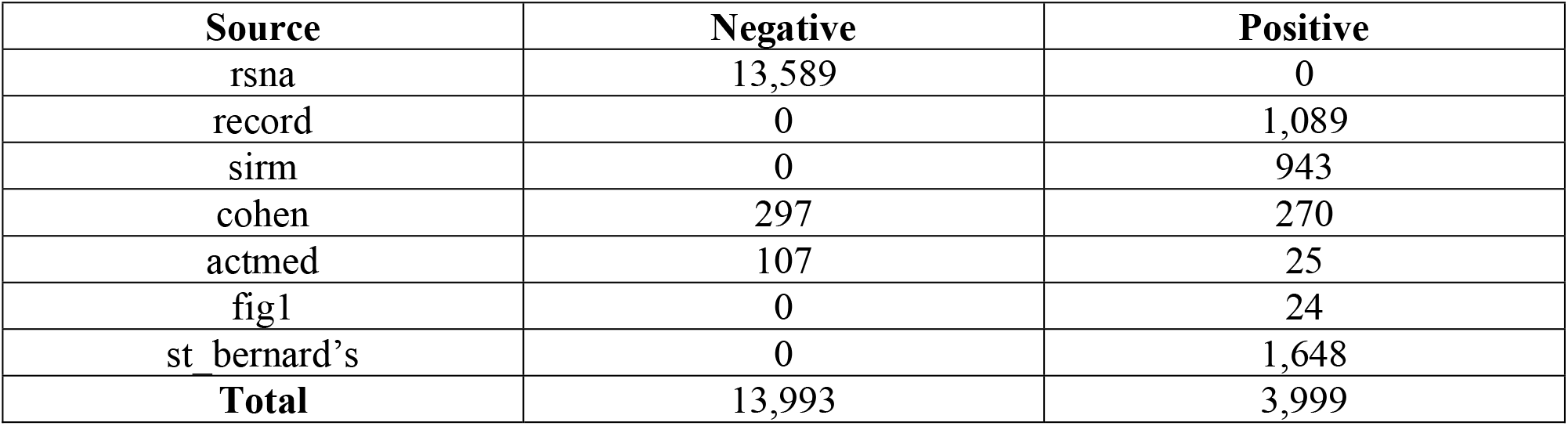

Despite having a larger dataset, it was still heavily imbalanced with approximately 3.5 times as many negative images than positive images. The St. Bernard’s dataset made up 41.2% of the positive samples in the dataset.

| Set | Negative | Positive |
| --- | --- | --- |
| Train | 11,022 | 2,787 |
| Valid | 2,771 | 682 |
| Test | 200 | 530 |

The original COVIDx dataset was split 80/20 between train/test sets. The test set was then split 80/20 again to create a validation set. 20% of the St. Bernard dataset was added to the test set and the rest was split 80/20 and added to the training and validation sets.

### Models to predict COVID-19 positive or negative with chest x-ray images

The dataset from St. Bernard’s was used to train a binary classifier to detect whether a given patient had Covid-19 or not. The COVIDx dataset contained chest x-rays for three classes: normal, pneumonia and, covid. Since this was a binary classification task the normal and pneumonia classes were combined into a single “negative” class and the covid images were the “positive” class. A DenseNet model was used as the classifier for this task using pretrained ImageNet weights. ImageNet is a dataset that contains images for 1,000 categories. We used transfer learning to apply a pretrained DenseNet model on the covid classification task.

The data was preprocessed into 224×224 images and central cropped. On each training epoch the images were shuffled and randomly augmented using rotations, translations, brightness adjustment, zoom, skewing and ratio resizing.

The model was then retrained to test generalizability of training a model on x-ray images from one hospital/clinic and then predicting on images captures at another. This was done by removing all the images from St. Bernard’s from the training set and using the same test set as the previous task so that they could be directly compared. This hold-one-out experiment was conducted to test the model’s generalizability.

### The performance of the prediction model and the generalizability

The model completed after 10 epochs of training and used the class weights to account for the class imbalance. The model attained an accuracy of 95.9%, precision of 98.1%, recall of 96.2% and an f1-score of 97.1%. Several models were trained with the training time varying between them; however, this model was selected as it had the fewest rate for false negatives at 2.74%. Given the severity of covid-19 at this time it was decided that a model that minimized false negatives was prioritized over one that simply maximized accuracy.

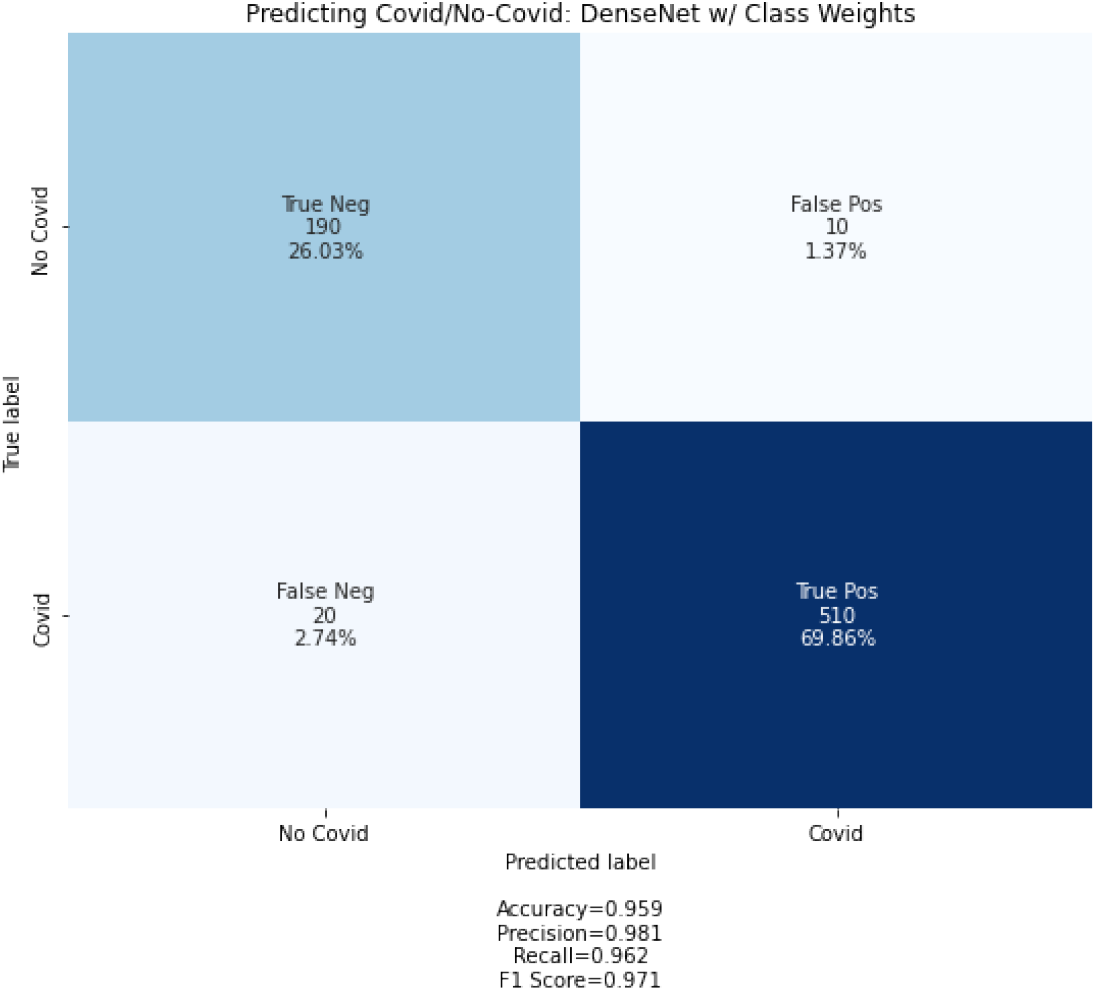

The model that was trained without using images from St. Bernard performed poorly during testing. It had an accuracy of 71.2%, precision of 100%, recall of 60.4% and an f1-score of 75.3%. This could be due to a combination of multiple reasons. First, the St. Bernard dataset accounted for 41.2% of all the positive samples so removing them would certainly have a significant impact on the model’s ability to learn positive cases. Second, this could also be due to the model’s inability to generalize to images from another hospital. There may be differences in the machines and techniques used that could hamper a model’s performance if it has not been trained on similar images. Given that 82.9% of the false negatives were from the St. Bernard dataset it does lend evidence to this being a problem for the model. It was not able to generalize to this new dataset.

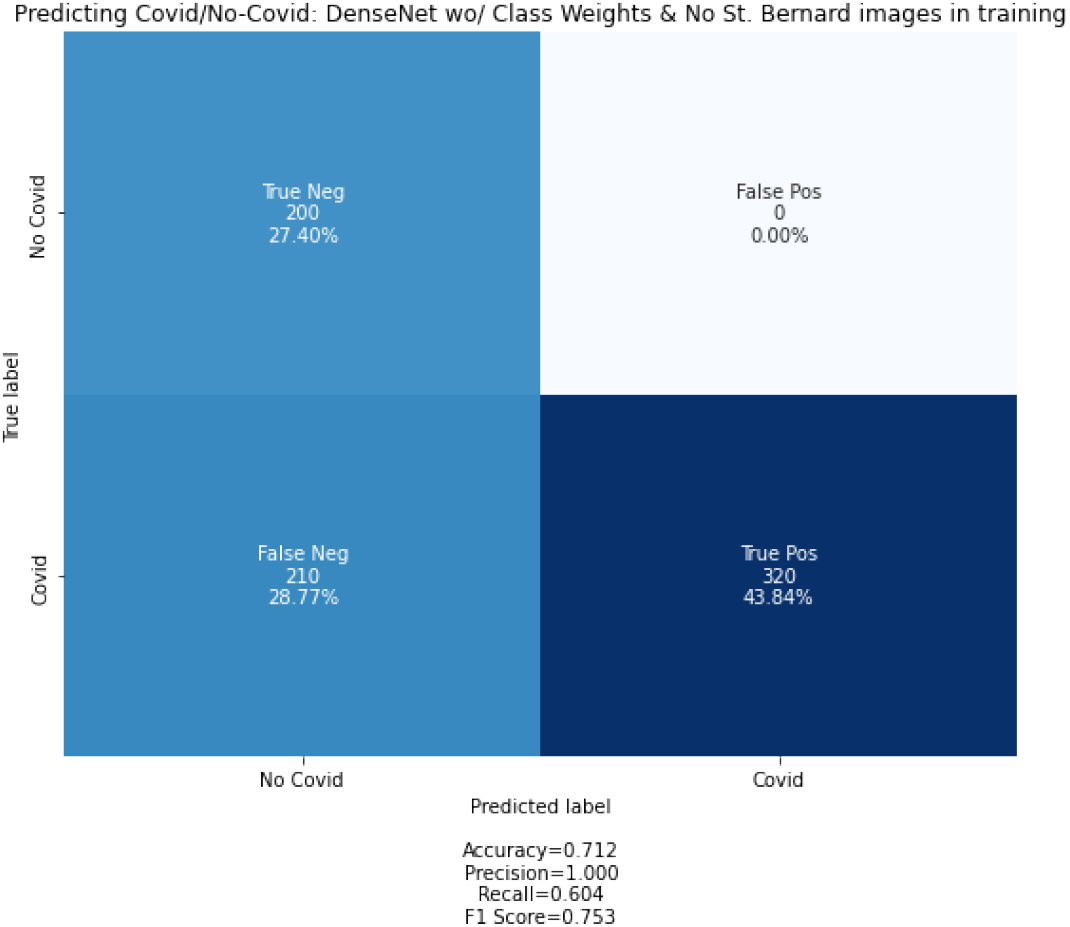

## Discussion of prediction improvements and future research

A significant hurdle for model performance was the imbalanced data set. As the dataset is expanded with either additional images or if the current dataset can be supplemented with clinical data, then we believe the models could overcome this issue. We also plan to use a model that has been trained with other COVID-19 chest x-rays for transfer learning. Although CheXnet was trained with chest x-rays, these were of other diseases and may not transfer as well to this task. By using a model pre-trained on other chest x-rays, it may extract features that are more relevant to predicting the duration of a patient’s hospital stay.

A larger and more diverse dataset would help with analysis in the binary classification tasks along with more samples of positive covid cases from a larger range of facilities. This would lessen the impact of a hold-one-out study of the generalizability of models trained on chest x-rays. This way we could repeat the hold-one-out experiments without the model suffering due to a significant reduction in training samples of any class.

## Data Availability

All data produced in the present study are available upon reasonable request to the authors

## ACKNOWLEDGMENTS

This research work and effort was partially supported by the National Science Foundation EPSCoR grant number 1946391, and the National Science Foundation with grant number 1239812, 1452211, 1553680, 1723529, 2054737, and 2216084, National Institute of Health grant R01LM012601, as well as was partially supported by National Institute of Health grant from the National Institute of General Medical Sciences (P20GM103429).

## Reference

Kenneth McIntosh, MD. Coronavirus disease 2019 (COVID-19): Epidemiology, virology, and prevention. (ed. Allyson Bloom, MD) (2020).

John Hopkins Coronavirus Resource Center, https://coronavirus.jhu.edu/map.html

Chinese Center for Disease Control and Prevention, https://www.chinacdc.cn/en/

Center for Disease Control and Prevention, https://www.cdc.gov/

Stubblefield J, Hervert M, Causey JL, Qualls JA, Dong W, Cai L, Fowler J, Bellis E, Walker K, Moore JH, Nehring S, Huang X. Transfer learning with chest X-rays for ER patient classification. Sci Rep. 2020 Dec 1;10(1):20900. doi: 10.1038/s41598-020-78060-4. PMID: 33262425; PMCID: PMC7708466.

Wang L, Lin ZQ, Wong A. COVID-Net: a tailored deep convolutional neural network design for detection of COVID-19 cases from chest X-ray images. Sci Rep. 2020 Nov 11;10(1):19549. doi: 10.1038/s41598-020-76550-z. PMID: 33177550; PMCID: PMC7658227.

Jenks, George F. 1967. “The Data Model Concept in Statistical Mapping”, International Yearbook of Cartography 7: 186–190.

McMaster, Robert, “In Memoriam: George F. Jenks (1916–1996)”. Cartography and Geographic Information Science. 24(1) p.56–59.

